# How AI is used in FDA-authorized medical devices: a taxonomy across 1,016 authorizations

**DOI:** 10.1101/2025.03.13.25323924

**Authors:** Rohan Singh, Monika Bapna, Abdul Rahman Diab, Emily S. Ruiz, William Lotter

**Author notes:** Correspondence to: William Lotter.

## Abstract

The recent proliferation of AI-enabled medical devices and the growing emphasis on clinical translation creates a critical need to understand the evolving scope of these technologies. We reviewed 1,016 FDA authorizations of AI/ML-enabled medical devices to develop a taxonomy that captures key axes of variation in clinical and AI-related features. While quantitative image analysis remains the most common application, the relative proportion of such devices has declined recently due to an increase in devices designed for different data types and higher-risk use cases. Notably, over 100 devices leverage AI for data generation, including synthetic image creation, though we did not currently find evidence of LLM-based generative models. Nonetheless, the FDA’s scope of AI/ML appears broad, encompassing devices that explicitly reference traditional ML methods such as K-nearest neighbors. To accompany the analysis, we have created a website to facilitate exploration of our curated database and trends over time. Altogether, our taxonomy and findings clarify how AI is currently used in medical devices and provide a foundation for tracking future developments as clinical applications evolve.

## Main

Over one thousand Artificial Intelligence (AI)/Machine Learning (ML)-enabled medical devices have been authorized by the U.S. Food and Drug Administration (FDA). Given the vast range of potential applications of AI in clinical care^1–5^ and the increasing focus on translating AI into routine practice, it is critical to understand what types of AI devices are currently authorized for clinical use, and how these use cases are evolving over time. The FDA’s current classification systems for medical devices, including product codes and device classes, provide a broad characterization that do not fully capture key dimensions of AI use. Recent work has provided insights into subsets of devices^6–11^, but a comprehensive characterization of FDA-authorized AI-enabled devices is currently lacking.

We reviewed all 1,016 authorizations of AI/ML-enabled medical devices listed by the FDA as of December 20th, 2024 and created a taxonomy to describe and quantify major axes of variation in current devices (**Fig. 1**). The taxonomy has three core factors. The first factor is the core *clinical function* of the device, describing the general medical role or purpose it serves in patient care.

**Figure 1:**
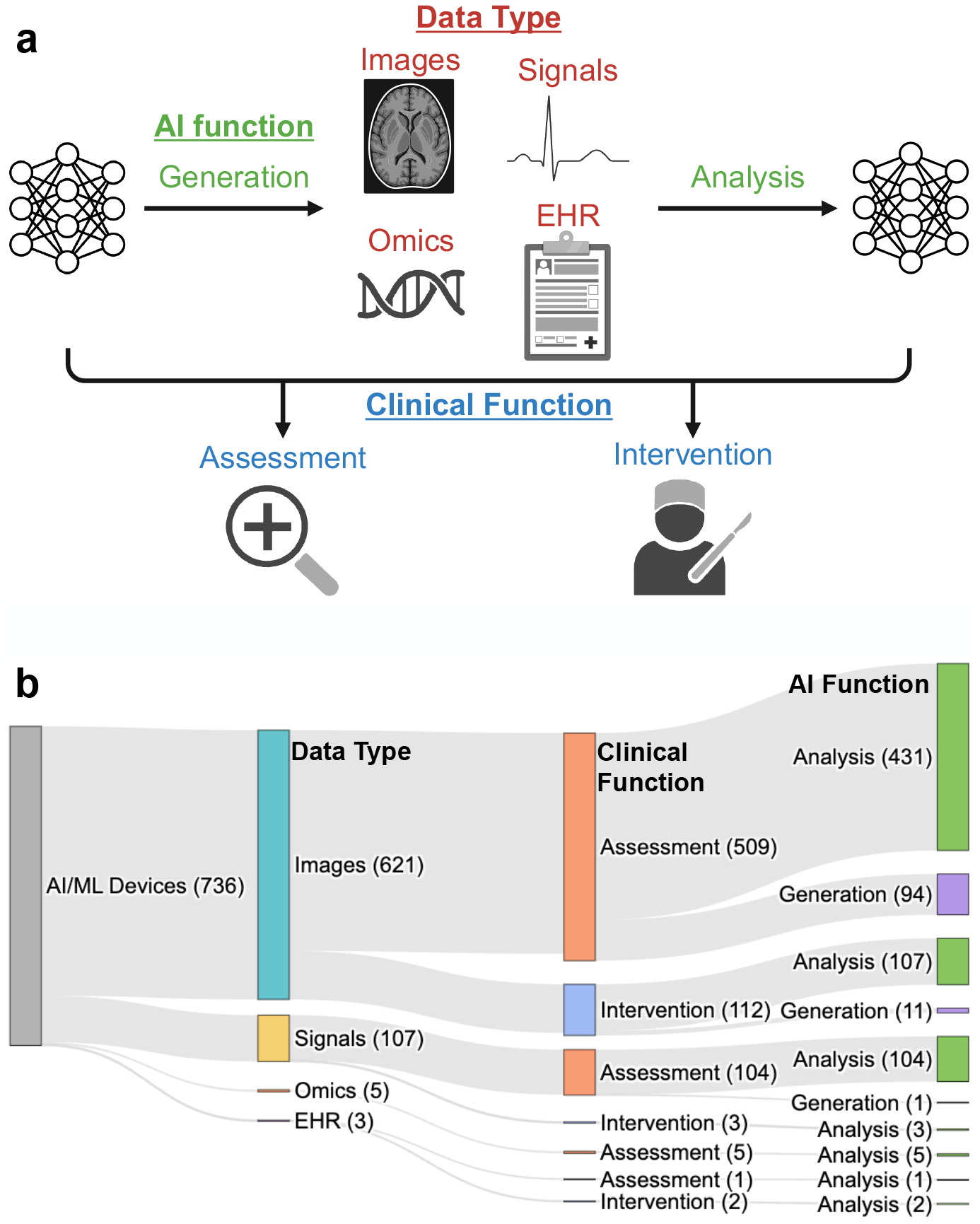
Taxonomy of AI/ML-enabled medical devices. a) Illustration of the three core factors: data type, AI function, and clinical function. b) Distribution of factor counts across FDA-authorized devices.

The second factor is the *AI function*, capturing how the device uses AI to assist with the clinical function. The third factor is the *data type* used as input to perform the AI function. By reviewing public authorization summaries for current devices, we categorized devices into different classes for each of these factors. As a given device can have multiple authorizations over time for updated versions, we additionally grouped the 1,016 authorizations into a list of unique devices through a semi-automated matching process (see Methods). We primarily report results at the device level rather than the authorization level to enable a more representative view of the current device distribution.

Across 1,016 FDA authorizations between 1995 and 2024, 736 unique devices were identified (**Supp. Table 1**). For data type, 621 (84.4%) devices use images as the core input to the AI algorithm, 107 (14.5%) use signals, 5 (0.7%) use ‘omics data, and 3 (0.4%) use tabular electronic health records (EHR). For devices based on imaging, Radiology was the lead review panel for the majority (88.2%), followed by Neurology (2.9%) and Hematology (1.9%). Signals include time series such as ECG and EEG, with Cardiovascular (64.5%), Neurology (16.8%), and Anesthesiology (12.1) representing the most common review panels. The five ‘omics-based devices use data pertaining to RNA expression, DNA variants, and/or antibody assays as input to the AI/ML model. The three current EHR-based devices use tabular data such as treatment information and vitals measurements as input.

In terms of clinical function, current devices fall into two broad categories based on whether they assist in patient assessment, such as diagnosis or monitoring, or intervention, such as surgery or treatment guidance. The majority of current devices fall under Assessment (84.1%). Of the 117 (15.9%) Intervention devices, 112 (95.7%) use images as the AI data type, followed by signals (3; 2.6%) and EHR (2; 1.7%). Current intervention applications for signals and EHR include assisting with insulin dosing based on either continuous glucose monitoring (signals) or tabular data (EHR), whereas common use cases for images include assisting with surgical or radiotherapy planning.

For AI function, we first classified devices based on whether the AI assists with the data generation and/or the data analysis process, as these categories often entail broadly different considerations for AI development and clinical implementation. We based this classification on the AI component of the device; for instance, an ultrasound machine that acquires images without using AI but has a built-in AI component to analyze the resulting images would be classified as “Analysis”. With this definition, we classified 630 (85.6%) devices as Analysis, 83 (11.3%) as Generation, and 23 (3.1%) devices use AI for both data generation and analysis. For the devices that use AI for data generation, 99% had a data type of Images.

Beyond Analysis vs. Generation, we subclassified the AI function of each device into more fine-grained categories based on the device outputs and intended use (**Fig. 2**). For Analysis, we identified 6 subclasses across current devices: triage, quantification/feature localization, detection, diagnosis, detection/diagnosis, and predictive. Triage devices output a binary prediction for a given exam or time series that serves as a notification for prioritized review by a clinician. Devices classified as quantification/feature localization calculate a quantitative metric from the input data and/or provide positional information of structural features. This was considered a joint category because these functions are often performed in tandem, such as segmenting an anatomical structure on a medical image and then quantifying its volume. Detection devices differ from quantification/feature localization in that they are specifically designed to assist clinicians in finding disease-suspicious regions in the input data, such as highlighting lung nodules on CT images or identifying the time interval of a potentially abnormal heart rhythm in an ECG trace. Devices indicated for diagnosis do not explicitly identify suspicious regions but instead output a score or category across the input data that indicates the likelihood of a specific diagnosis. Some devices assist with both detection and diagnosis, and we retain this as a distinct subclass as it is often considered as such in FDA regulation. Finally, predictive devices were defined as those that generate a score or category to indicate the future risk of an event or disease rather than the current risk.

**Figure 2:**
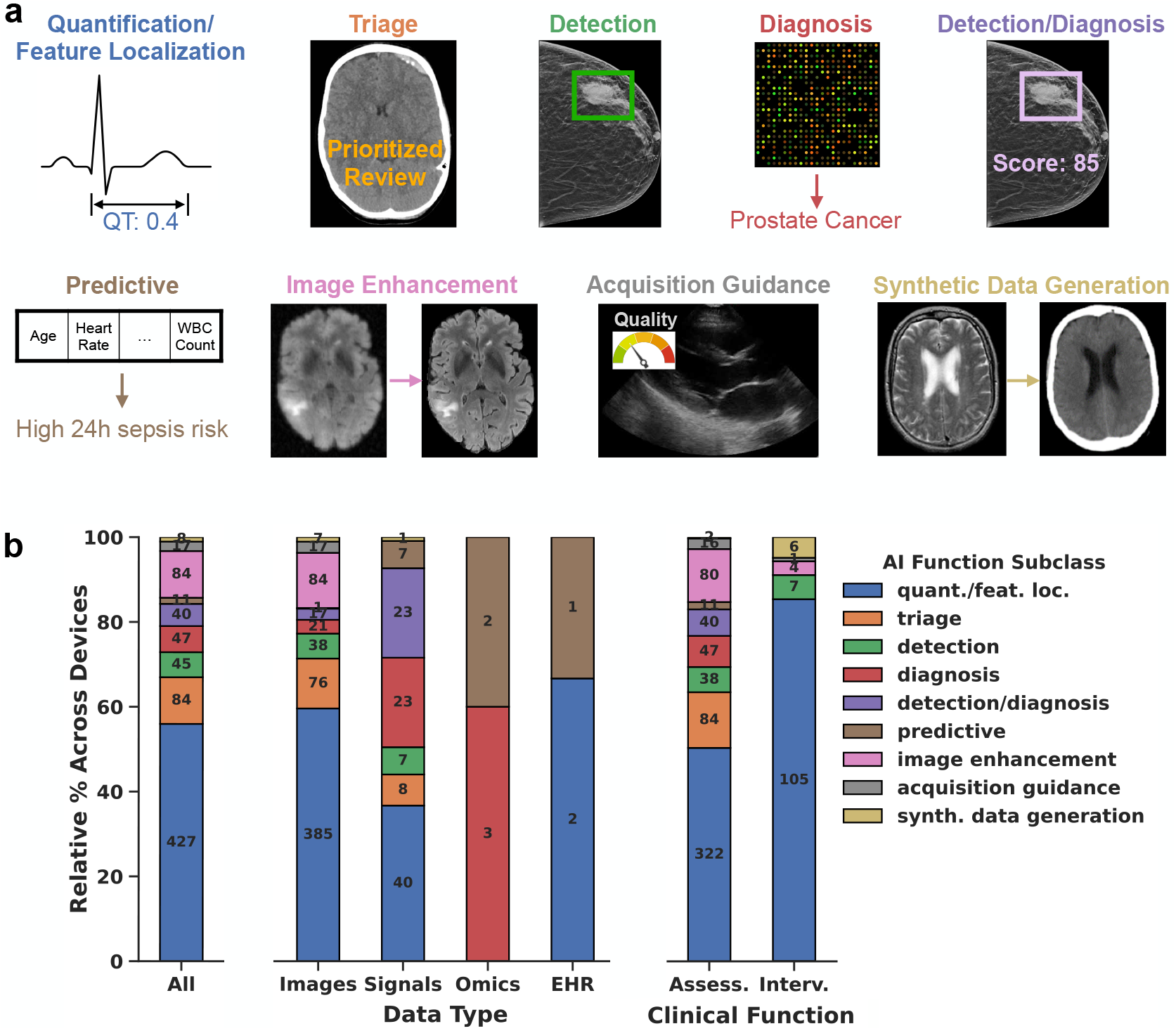
Subclasses of AI function in FDA-authorized medical devices. a) Illustration of the nine subclasses. b) Distribution of AI function subclasses across FDA-authorized devices. Counts indicate the number of devices with the AI function subclass.

Across the 653 Analysis devices, 427 (65%; 58% of all devices) were subclassified as performing quantification/feature localization. Triage was the next most common Analysis subclass with 84 devices (12.9% of Analysis devices; 11.4% of all devices). The remaining subclass counts were as follows: 47 (7.2/6.4%) diagnosis, 45 (6.9/6.1%) detection, 40 (6.1/5.4%) detection/diagnosis, and 11 (1.7/1.5%) predictive. The proportion of devices in each Analysis subclass varied by data type and clinical function (**Fig. 2**). Image-based devices have a relatively higher percentage of quantification/feature localization and triage, whereas signals-based devices have a relatively higher percentage of diagnosis, detection/diagnosis, and predictive.

For Generation devices, we identified 3 current subclasses: image enhancement, acquisition guidance, and synthetic data generation. Image enhancement includes tasks such as image denoising and AI-based reconstruction. Devices that use AI for acquisition guidance assist the clinical user in acquiring the appropriate data, such as indicating whether a body part or acquisition device is properly positioned. These devices do not explicitly generate the primary data used for downstream analysis but instead assist in properly capturing the data. Finally, synthetic data generation uses AI to generate a new data element altogether, such as synthesizing a CT medical image from MRI or synthesizing a particular ECG bandwidth lead from existing leads.

Of the 106 Generation devices, 84 (79.2%; 11.4% of all devices) perform image enhancement. Acquisition guidance and synthetic data generation included 17 (16.0/2.3%) and 8 (7.5/1.1%) devices, respectively. Images were the data type for 105 of the 106 Generation devices, with the remaining device performing synthetic data generation using signals.

In addition to quantifying the current distribution of AI-enabled devices, we explored trends over time. While Images remains the most common data type class, its relative proportion among new devices peaked in 2021 at 94% and stands at 81% in 2024 so far (**Fig. 3**). Similarly, quantification/feature localization is the most common AI function subclass, but its prevalence peaked at 81% of devices in 2016 and has declined to 51% in 2024. The proportion of triage and image enhancement especially increased between 2017 and 2021, with 2024 now containing a relatively mixed proportion of devices across the subclasses. For clinical function, the ratio of Assessment vs. Intervention has remained relatively consistent over time. These trends also hold at the individual authorization level rather than the device level (**Supp. Fig. 2, Supp. Fig. 3**). The percentage of authorizations that represent updated versions of existing devices has averaged 34% over 2022-2024, compared to 14% between 2017-2019 (**Supp. Fig. 1**).

**Figure 3:**
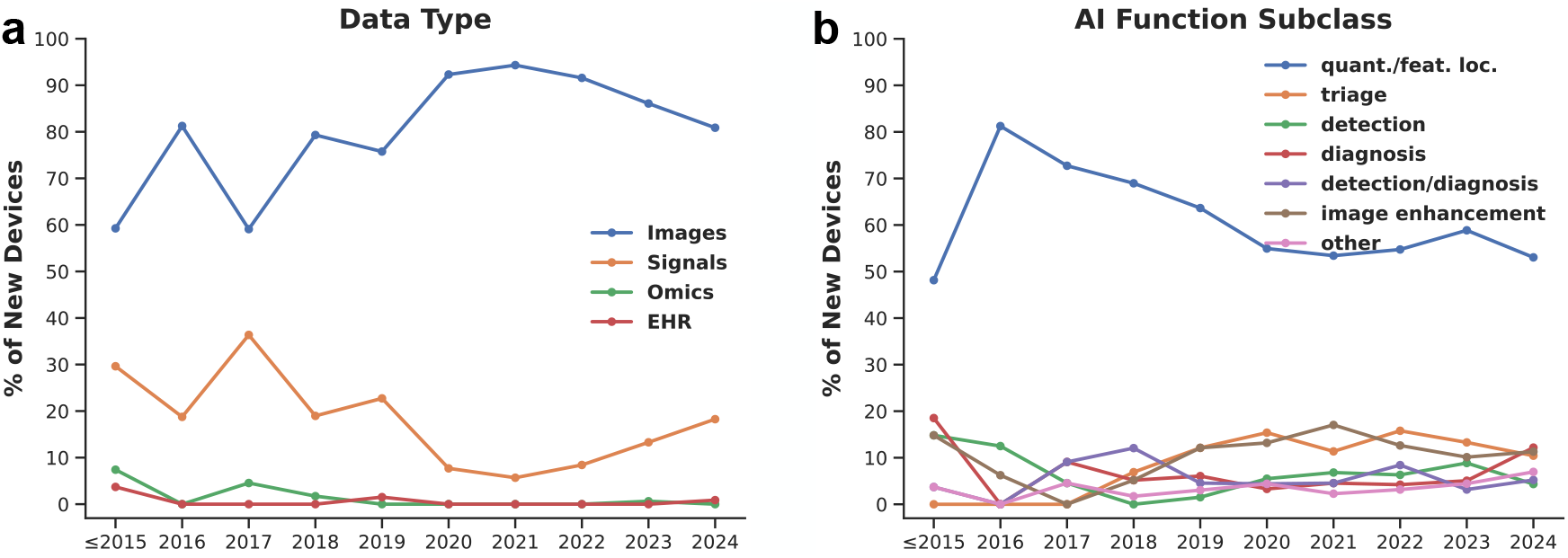
Trends in data type and AI function subclass over time. The percentage of new devices per year is displayed by a) data type and b) AI function subclass. The AI function subclasses of predictive, synthetic data generation, and acquisition guidance are grouped as “other” given low counts.

Our review of the 1,016 authorizations revealed several other notable insights. First, while the specific AI/ML methods used by a device were often unclear, it is evident that the FDA employs a broad definition of AI/ML in its current device list. For instance, some devices explicitly refer to the use of traditional (non-deep learning) ML methods in their authorization summaries. In fact, the NVI product code, which currently includes one ‘omics-based device, includes the K-nearest neighbors algorithm in its name: “Diagnostic Software, K-Nearest Neighbor Algorithm, Autoimmune Disease”. When deep learning was used, certain devices indicated the specific model and/or architecture family, such as convolutional or recurrent neural networks. However, we did not find evidence of large language models (LLMs) in the studied device list.

Along with a common lack of clarity regarding the form of AI/ML, it was unclear where AI/ML was used altogether in some authorization summaries. This ambiguity was particularly common for devices that perform many functions, some of which may or may not involve AI/ML. In such cases, we estimated the taxonomy classes based on similar devices, domain expertise, and/or available information from manufacturer websites. As a result, our findings should be interpreted in terms of overall trends rather than precise class counts. Another limitation of our study is its reliance on manual curation (see Methods). While we explored various LLMs and prompting strategies for automated classification, these methods proved insufficient given the nuances and variability of the data.

Rather than treating AI as a homogeneous technology in medicine, it is essential to understand current use cases and evolving trends. We created an organized taxonomy of AI/ML-enabled medical devices by reviewing over one thousand FDA authorizations. Our analysis confirms some expected patterns, such as a high proportion of quantitative imaging devices, but also highlights notable recent shifts, such as increases in higher risk use cases and signals-based devices. While LLM-based generative models are not yet represented, more than one hundred devices leverage AI for medical data generation, including image denoising (“super-resolution”^12^) and synthetic data creation. To facilitate further exploration, we have developed an interactive website (https://fda-ai-taxonomy.vercel.app/) to host the curated database. Our taxonomy is designed not only to capture the current landscape but also to be extendable as new devices receive authorization. Continued monitoring of AI-enabled medical devices will be crucial for healthcare professionals, policymakers, and AI developers to fully realize AI’s translational potential in medicine.

## Methods

### Database curation process

To create the database, the FDA’s published list of AI/ML-enabled devices (https://www.fda.gov/medical-devices/software-medical-device-samd/artificial-intelligence-and-machine-learning-aiml-enabled-medical-devices) was analyzed. The version accessed corresponded to an update on December 20th, 2024, which included devices authorized up to September 27th, 2024. From this list, we curated the taxonomy over two phases. First, we reviewed all of the unique product codes across the devices, including reviewing the description and example authorizations for each code. The descriptions are available via the FDA’s Product Code Classification database and authorization summaries were accessed using the FDA’s 510(k), De-novo, and PMA databases. For each product code, we determined whether the taxonomy values could potentially vary across devices within the product code, or whether all devices would clearly have the same values. For example, the QFM product code has a description of “Radiological computer aided triage and notification software”, which would correspond to a data type of Images and AI function subclass of triage for all devices with this code. Every product code was reviewed by at least two members of the study team. Any product code for which any of the taxonomy levels could potentially vary by device were then considered for the second phase of review, which involved reviewing the individual authorization summaries for each device within the product code. This review was performed by the senior author.

### Taxonomy assignments

Each device was classified according to four factors: data type, clinical function, AI function, and AI function subclass. The possible classes for each factor were defined through the review process to encompass major axes of variation across the devices, and were also informed by existing categories of medical devices^7,8^, such as computer-aided detection (“CADe”) and computer-aided diagnosis (“CADx”). Data type was assigned based on the core input to the AI algorithm in a mutually exclusive manner and could differ from the underlying data modality in some circumstances, as our goal was to characterize how AI is actually being used. For instance, if the device involved a chemical assay but only involved AI to quantify the results based on a picture of the assay acquired using a smartphone camera, the data type would be classified as “Images”. Clinical function was assigned based on the primary use case for the device according to the indications for use in the authorization summary. For imaging systems such as MRI machines that could theoretically be used for both diagnostic and interventional tasks, we considered the clinical use to be “Assessment”, unless the AI component was specifically designed to assist in interventions. Analogously, we assigned the AI function based on the output of the AI model(s) rather than the overall output(s) of the device. If a device had several AI-enabled components, we assigned the AI function for the device as a union across the individual components (i.e., a device could perform both data generation and analysis using AI).

### Determining unique devices

Along with performing the taxonomy assignment for each individual authorization, we grouped authorizations into a list of unique devices, since a given device can have multiple submissions over time (i.e., for updated versions). Authorizations were grouped as the same device if they had matching product codes and similar company names, device names, and indications for use. These groupings were first proposed automatically based on case-insensitive matching of product codes, company names, and device names. The authorization list and proposed matches were then manually reviewed to make any corrections, including comparing the authorization summaries across potential matches to ensure similar indications for use when confirming matches, and separating preliminary matches that had the same device name but different indications for use. The latter scenario can occur when the device name on the submission represents a platform product, but the individual submissions pertain to unique AI algorithms (e.g., for detecting different diseases), in which case we considered each submission unique. The manuscript includes results presented at both the authorization and device level. When presenting results by device, we use the most recent authorization unless otherwise noted.

### Trend analysis

When performing the trend analysis at the device level, the earliest authorization for each device was considered. Taxonomy counts per year were analyzed, where devices/authorizations before 2016 were grouped together given low counts per year (33 total authorizations between 1995 and 2015 with a maximum of 6 per year over this timeframe).

## Data Availability

The curated taxonomy database is available as Supplementary Data 1.

**Supplementary Table 1:**
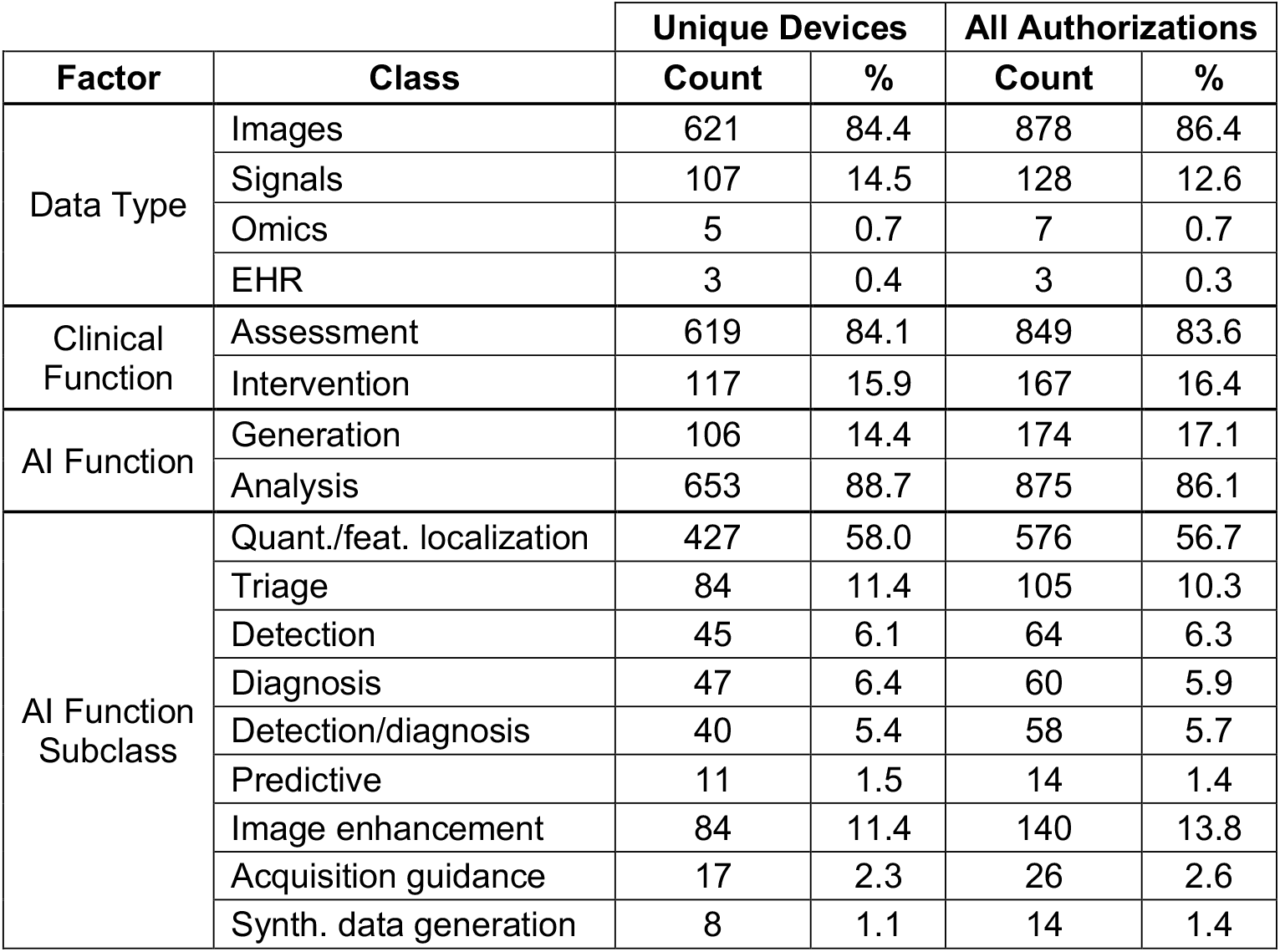
Counts of devices and authorizations across all taxonomy factors.

**Supplementary Figure 1:**
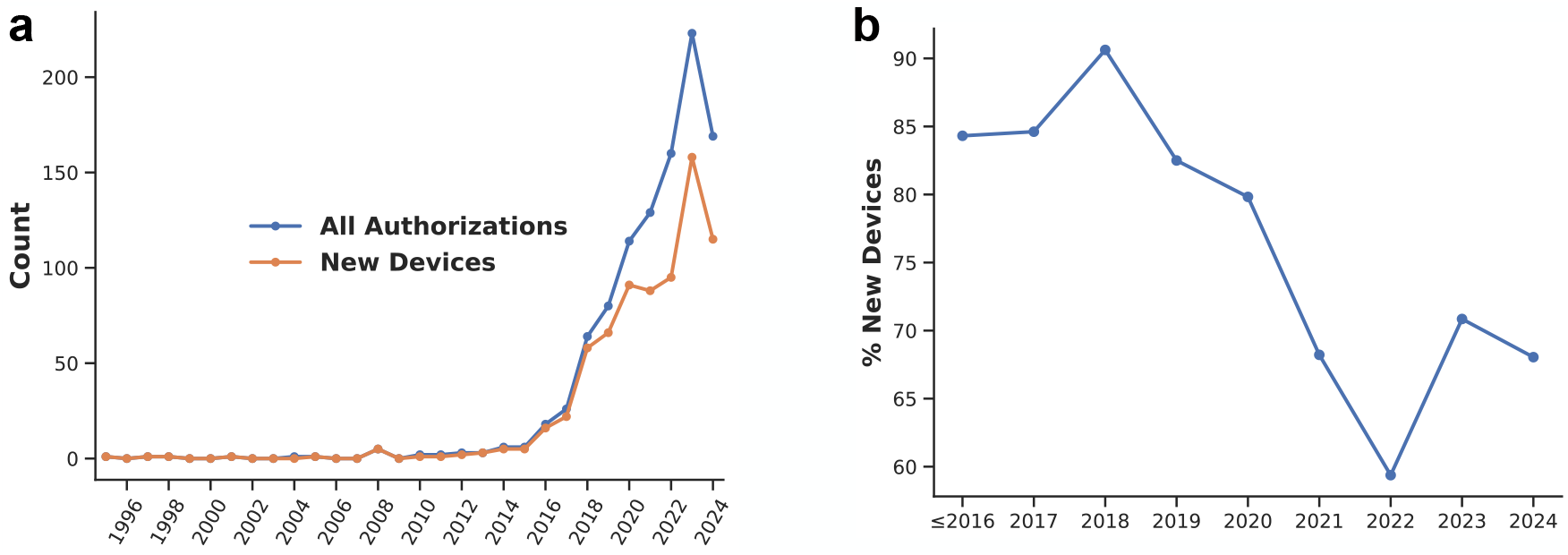
Number of authorizations and new devices over time. a) Counts from 1995 to 2024 (up to September 27th). b) Percentage of authorizations that represent new devices.

**Supplementary Figure 2:**
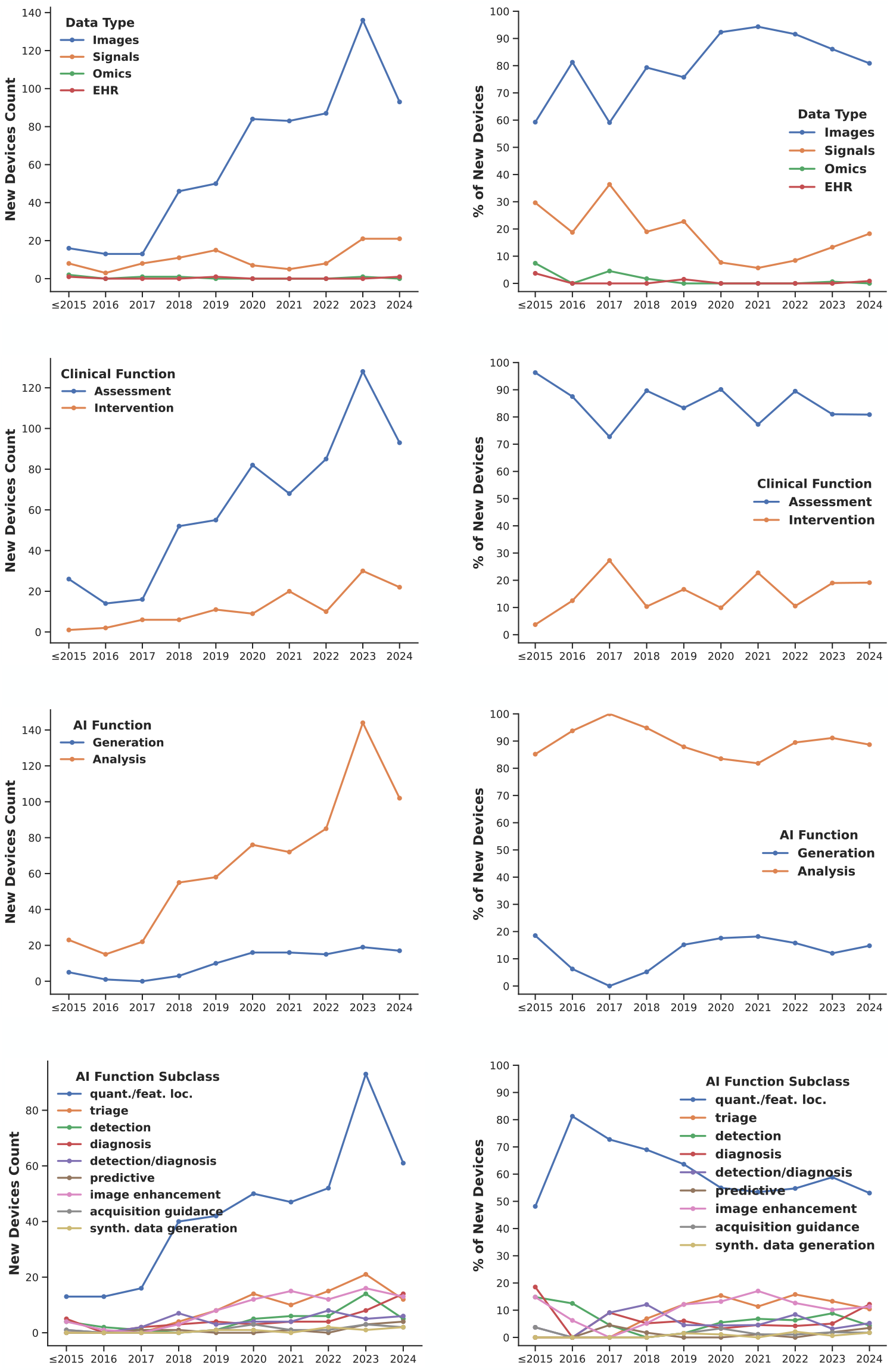
Counts and percentages of new devices per year for each of the taxonomy factors.

**Supplementary Figure 3:**
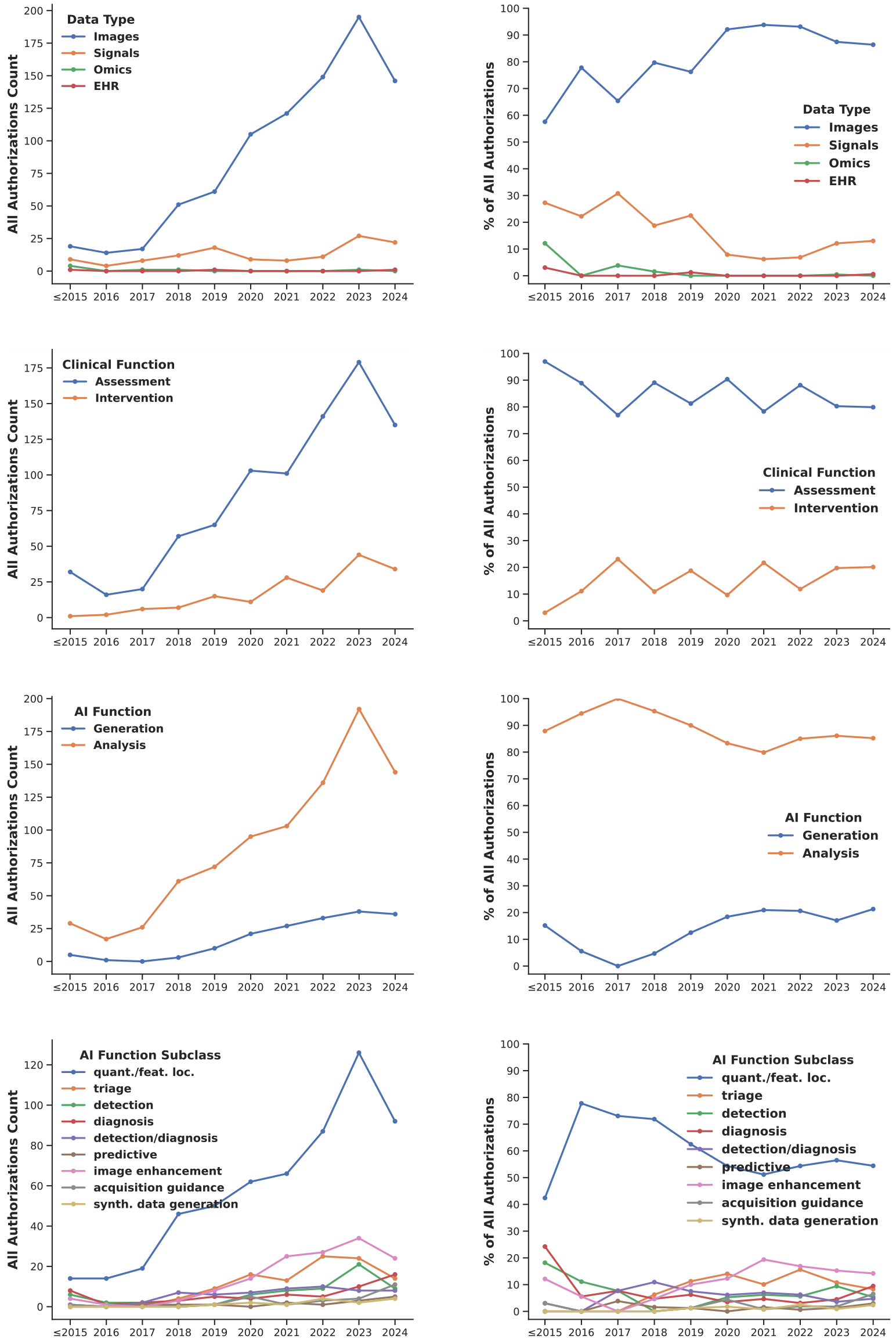
Counts and percentages of all authorizations per year for each of the taxonomy factors.

